# Humoral immune responses against SARS-CoV-2 variants including omicron in solid organ transplant recipients after three doses of a COVID-19 mRNA vaccine

**DOI:** 10.1101/2021.12.29.21268529

**Authors:** Kapil K. Saharia, Jennifer S. Husson, Silke V. Niederhaus, Thierry Iraguha, Stephanie V. Avila, Youngchae J. Yoo, Nancy M. Hardy, Xiaoxuan Fan, Destiny Omili, Alice Crane, Amber Carrier, Wen Y. Xie, Erica Vander Mause, Kim Hankey, Sheri Bauman, Patricia Lesho, Heather D. Mannuel, Ashish Ahuja, Minu Mathew, James Avruch, John Baddley, Olga Goloubeva, Kirti Shetty, Saurabh Dahiya, Aaron P. Rapoport, Tim Luetkens, Djordje Atanackovic

## Abstract

**Background:** Solid organ transplant recipients (SOTR), who typically receive post-transplant immunosuppression, show increased COVID-19-related mortality. It is unclear whether an additional dose of COVID-19 vaccines in SOTR can overcome the reduced immune responsiveness against Severe Acute Respiratory Syndrome Coronavirus 2 (SARS-CoV-2) variants.

**Methods:** We performed a prospective cohort study of 53 SOTR receiving SARS-CoV-2 vaccination into a prospective cohort study performing detailed immunoprofiling of humoral immune responses against SARS-CoV-2 and its variants.

**Results:** Prior to the additional vaccine dose, 60.3% of SOTR showed no measurable neutralization and only 18.9% demonstrated neutralizing activity of >90% following two vaccine doses. More intensive immunosuppression, antimetabolites in particular, negatively impacted antiviral immunity. While absolute IgG levels were lower in SOTR than controls, antibody titers against microbial recall antigens were in fact higher. In contrast, SOTR showed reduced vaccine-induced IgG/IgA antibody titers against SARS-CoV-2 and its delta variants. Vaccinated SOTR showed a markedly fewer linear B cell epitopes, indicating reduced B cell diversity. Importantly, a third vaccine dose led to an increase in anti-SARS-CoV-2 antibody titers and neutralizing activity across alpha, beta and delta variants. However, we observed a significant decrease in anti-spike antibody titers with the omicron variant.

**Conclusions:** Only a small subgroup of SOTR generated functionally relevant antibodies after completing the initial vaccine series based on dysfunctional priming of immune responses against novel antigens. An additional dose of the vaccine results in dramatically improved antibody responses against all SARS-CoV-2 variants except omicron.

## INTRODUCTION

COVID-19 is caused by Severe Acute Respiratory Syndrome Coronavirus 2 (SARS-CoV-2) [1] which contains several structural proteins including the surface-exposed spike (S) and the internal nucleocapsid (N) proteins [1, 2]. The S fusion protein consists of the S1/S2 components and the virus enters cells, such as pneumocytes in the lung [3], through binding of the receptor-binding domain (RBD) within the S1 protein [4], to the angiotensin-converting enzyme-2 (ACE-2) receptor [2, 5]. Older patients and patients with pre-existing medical conditions, including different types of cancer [6], show a more complicated course of COVID-19 and have a worse prognosis [7-10]. Solid organ transplant recipients (SOTR) with COVID-19 also show an increased mortality of >20% [11-17]. In these patients, advanced age and comorbidities such as cardiovascular and pulmonary disease seem to contribute to the reduced survival [18-20].

The development of anti-SARS-CoV-2 antibodies following an active infection and/or vaccination, especially those directed against the S/RBD proteins of the virus, is crucial for the (1) protection from future COVID-19 infections, (2) limiting disease severity, and (3) controlling viral transmission [21, 22]. Unfortunately, SOTR receive long-term immunosuppressive treatment and are, therefore, less likely to build a protective immune response against SARS-CoV-2 [23-26]. Indeed, it was shown that following an active COVID-19 infection only ∼50% of SOTR will mount an antibody response [27] with kidney transplants vs. other types of transplants, shorter time from transplant to diagnosis, and more intensive immunosuppression being associated with a reduced humoral immune response [27].

The U.S. Food and Drug Administration (FDA) has issued emergency use authorizations or full approval for 3 vaccines for the prevention of COVID-19 [28, 29]. All three vaccines have been shown to elicit antibody- and T cell-mediated antiviral immune responses which confer almost complete protection against infection with the COVID-19 virus in healthy individuals [30-36]. Unfortunately, recent studies have indicated that SOTR, in agreement with observations made after an active COVID-19 infection, show reduced antibody responses following COVID-19 vaccination [37-40] against the original SARS-CoV-2 virus.

Until very recently, the more infectious B.1.617.2 (delta) variant of SARS-CoV-2 contributed to a surge in cases across the globe [41]. Only modest differences in vaccine effectiveness were noted with the delta variant compared to the original viral strain following two doses of COVID-19 mRNA vaccines, however, this may be very different in immunocompromised individuals, such as SOTR [42]. Additionally, the recently spreading omicron variant may even further promote viral immune escape [43] in these patients. Unfortunately, a detailed picture of vaccine-induced antibody responses in SOTR, including a description of immunodominant antibody epitopes, has never been obtained. Furthermore, it has remained unclear whether the administration of an additional dose of the vaccine can overcome the reduced immune responsiveness in SOTR especially against the different variants of SARS-CoV-2 such as the delta and the omicron mutants. Here, we present the results of our prospective study investigating in detail humoral immune responses against SARS-CoV-2 and its recent variants in fully vaccinated SOTR and in response to an additional dose of SARS-CoV-2 vaccine.

## METHODS

### Study Population and Design

We performed a prospective cohort study of SOTR 18 years of age or older who received post-transplant care at the University of Maryland Medical Center (UMMC) and had either received or were scheduled to receive any of the three SARS-CoV-2 vaccines approved by the FDA under emergency use authorization. Those who were already vaccinated must have received their initial vaccine dose within approximately 90 days of study enrollment. SARS-CoV-2 vaccination was not provided as part of this study protocol. Patients were excluded if they were HIV positive, were receiving chemotherapy or radiation therapy, or previously had SARS-CoV-2 infection. Demographic and clinical data were collected from study participants and through medical chart abstractions. Up to 50 ml of heparinized blood was collected at the following time points (see Supplemental Fiigure 1): visit 1 (within 7 days prior, up to 48 hours after first vaccine dose), visit 2 (+/- 7 days), visit 3 (at 3 months [+/- 10 days] after 1st vaccine dose), visit 4 (at 6 months [+/- 14 days] after 1st vaccine dose), visit 5 (at 9 months [+/- 14 days] after 1st vaccine dose), and visit 6 (at 12 months [+/- 14 days] after 1st vaccine dose). All patients who received a mRNA COVID-19 vaccine received the second dose 3-4 weeks after the first dose. For the post-third dose analysis, samples were chosen from the available samples at least two weeks after the third dose was administered. Plasma was generated from peripheral blood samples after centrifugation at 400g for 10min and frozen immediately at -70C. Peripheral blood mononuclear cells (PBMCs) were isolated using lymphocyte separation density gradient and immediately frozen in liquid nitrogen. This study was reviewed and approved by the University of Maryland (Baltimore) institutional review board (HP-00095043).

For the epitope screening, samples from patients with an active COVID-19 infection were used. These samples were collected as part of our prospective observational study enrolling COVID-19 patients who were admitted to the University of Maryland Medical Center between June and August of 2020. For that study, informed consent was obtained and blood samples were collected under IRB HP-00091425. Samples from vaccinated healthy controls were collected as part of our prospective clinical study on immune responses to two doses of a COVID-19 mRNA vaccine in cancer patients and healthy controls (IRB HP-00095016).

### Measurement of absolute immunoglobulin levels

Absolute serum concentrations of the different immunoglobulins were measured using Human IgG, IgM, and IgA Enzyme-linked Immunosorbent Assay (ELISA) Kits (Invitrogen, Cat. No. BMS2091, BMS2098, BMS2096) as per the manufacturer’s instructions. Absorbance was read at 450nm with a reference wavelength of 620nm in a microtiter plate reader (Tecan, Morrisville, NC).

### Analysis of SARS-CoV-2-specific antibodies

Serum antibody responses against recombinant, full-length SARS-CoV-2 proteins (Supplemental Table 2), viral control proteins (Supplemental Table 1), or overlapping peptides covering the complete amino acid sequence of the SARS-CoV-2 S1 protein were determined by ELISA as previously described [44-46]. Briefly, high-binding ELISA plates (Thermo Fisher, Cat. No. 44-2404-21) were coated with 5µg/mL of the respective proteins in PBS (Gibco, Cat. No. 10010-023) overnight at 4°C. The next day plates were washed twice with PBS and twice with 0.1% PBS-T (VWR, Cat. No. M147-1L). Plates were then blocked with 5% non-fat dry milk (Santa Cruz, Cat. No. sc2325) in PBS (MPBS) for 1h at room temperature (RT), then washed again as described above. Serum was diluted 1:40 for screening assays and for titration 1:100/1:400/1:1,600/1:6,400 and if necessary 1:25,000 and 1:100,000 in MPBS. Diluted sera were added to plates and incubated for 3H at RT. Plates were washed as described above before incubation with secondary antibodies against pan-human IgG (Southern Biotech, Cat. No. 2040-04) or IgA (Southern Biotech, Cat. No. 2050-04). Secondary antibodies were diluted according to the manufacturers’ instructions and plates incubated for 1h at RT. Plates were then washed as described above, PNPP tablets (Southern Biotech, Cat. No. 0201-01) dissolved in diethanolamine (Thermo, Cat. No. 34064) and PNPP substrate solution added to each well for 10min in the dark. 15uL of 3N NaOH (VWR, Cat. No. BDH7472-1) stop solution was added to each well and absorbance was read at 405nm with a reference wavelength of 620nm in a microtiter plate reader (Tecan, Morrisville, NC). Endpoint titers were calculated using serum titration curves for positive samples and pooled sera of 5 healthy donors. For non-SARS-CoV-2 antigens, serum dilutions for anti-GST (glutathione-S-transferase) antibodies (Supplemental Table 1) were used as a negative control.

For peptide ELISAs, plates were first coated with 5ug/ml neutravidin (Thermo, Cat. No. 31000) overnight at 4°C and then blocked with 2% bovine serum albumin (BSA; Thermo, Cat. No. 9048-46-8) in PBS for 1h at RT. Plates were then incubated for 1h at RT with either 1ug/mL of the individual peptides or 5ug/mL equimolar peptide pools in PBS as indicated. Plates were washed and then developed with serum at a dilution of 1:40 and with secondary reagents as described above using 2% BSA instead of M-PBS.

### SARS-CoV-2 neutralization assay

Neutralizing activity of patient sera was assessed using the cPass Neutralization Antibody Detection Kit (GenScript, Cat. No. L00847-A) which is a surrogate test detecting circulating neutralizing antibodies against SARS-CoV-2 that block the interaction between the receptor binding domain (RBD) of the viral spike glycoprotein with the ACE2 cell surface receptor. Briefly, samples and controls were diluted with sample dilution buffer and pre-incubated with the Horseradish peroxidase (HRP) conjugated recombinant SARS-CoV-2 RBD fragment (HRP-RBD) or one of its variants listed in Supplemental Table 2 to allow the binding of the circulating neutralization antibodies to HRP-RBD. The mixture was then added to the capture plate, which was pre-coated with the hACE2 protein. The unbound HRP-RBD as well as any HRP-RBD bound to non-neutralizing antibody was captured on the plate, while the circulating neutralization antibodies HRP-RBD complexes remained in the supernatant and were removed during washing. Following a wash cycle, TMB substrate solution was added followed by the Stop Solution. The absorbance of the final solution was read at 450 nm in a microtiter plate reader (Tecan, Morrisville, NC).

### Statistical analyses

Statistical analyses for serological analyses were performed using GraphPad Prism 9 software (GraphPad Software, San Diego, CA). Groups were compared using the Mann–Whitney U test and paired analyses were performed using the Wilcoxon signed-rank test. Correlations were calculated using the Pearson correlation coefficient. For the analysis of clinical characteristics, groups were compared using a student’s t-test.

## RESULTS

### Solid organ transplant recipients show a markedly reduced antibody-mediated neutralizing activity in response to mRNA COVID-19 vaccines

As a first step, we screened a total of 53 SOTR recipients and 5 healthy controls for neutralizing antibodies inhibiting RBD-ACE2 interactions at 8 weeks after receiving the second dose of an mRNA COVID-19 vaccine or 90 days after receiving the J&J vaccine. We found that all healthy controls (Figure 1) showed a viral neutralization >90% and were therefore classified as “Good Responders” (GR). In marked contrast, most of the SOTR (32/53; 60.3%) showed a viral neutralization of less than 30% and were therefore classified as “Non-Responders” (NR). Of the 53 SOTR, 11 (20.8%) showed a neutralizing activity between 30% and 90% and were classified as “Reduced Responders” (RR). Only 10 (18.9%) showed a neutralizing activity of above 90% and were classified as GR. (Figure 1). Overall, the vast majority of SOTR (81.1%) showed a neutralizing activity <90% after completing the initial vaccine series.

**Figure 1:**
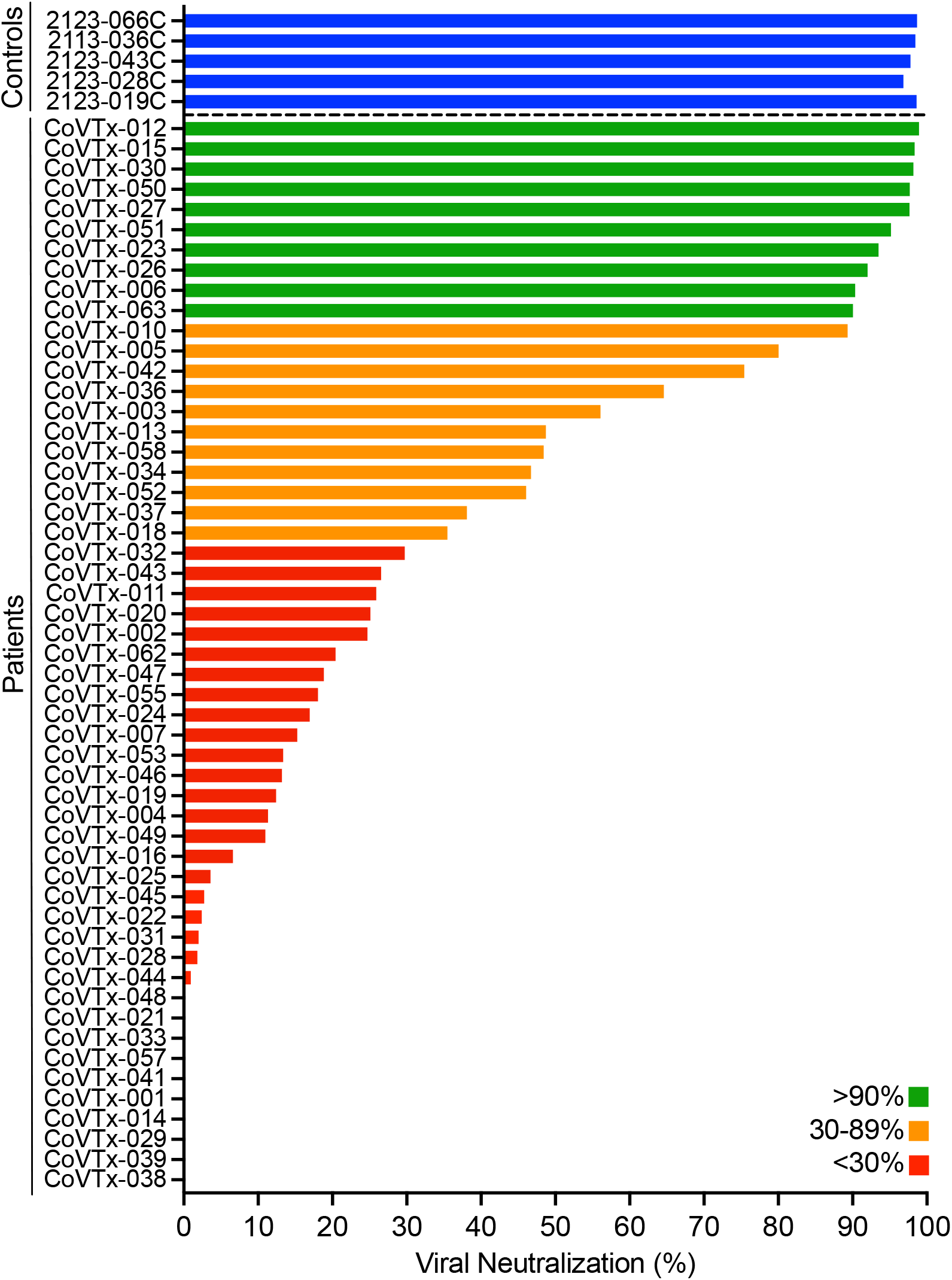
Neutralizing activity in the peripheral blood of SOT recipients after two doses of a COVID-19 mRNA vaccine. Neutralizing activity of vaccine-induced anti-RBD antibodies in the peripheral blood of SOT recipients (N=53) and healthy controls (N=5; blue bars) after the second dose of the vaccine was measured as the degree of inhibition of RBD-ACE2 interactions. Green, orange, and red bars indicate different degrees of inhibition as indicated in the legend.

We did not find any significant associations (Table 1) of the patients’ demographic characteristics and their past medical history with the type of humoral immune response (GR vs. RR/NR). However, when we analyzed the patients’ clinical characteristics at the time of the first dose of the vaccine (Table 2), we found that a higher overall number of immunosuppressants was associated with a reduced vaccine-induced SARS-CoV-2 neutralizing activity. Importantly, among immunosuppressants, only treatment with antimetabolites, which prevent lymphocyte proliferation, had a significant negative impact on the patients’ antiviral humoral immunity (Table 2), indicating that specifically antimetabolites interfere with the development of COVID-19-specific antibodies.

**Table 1:**
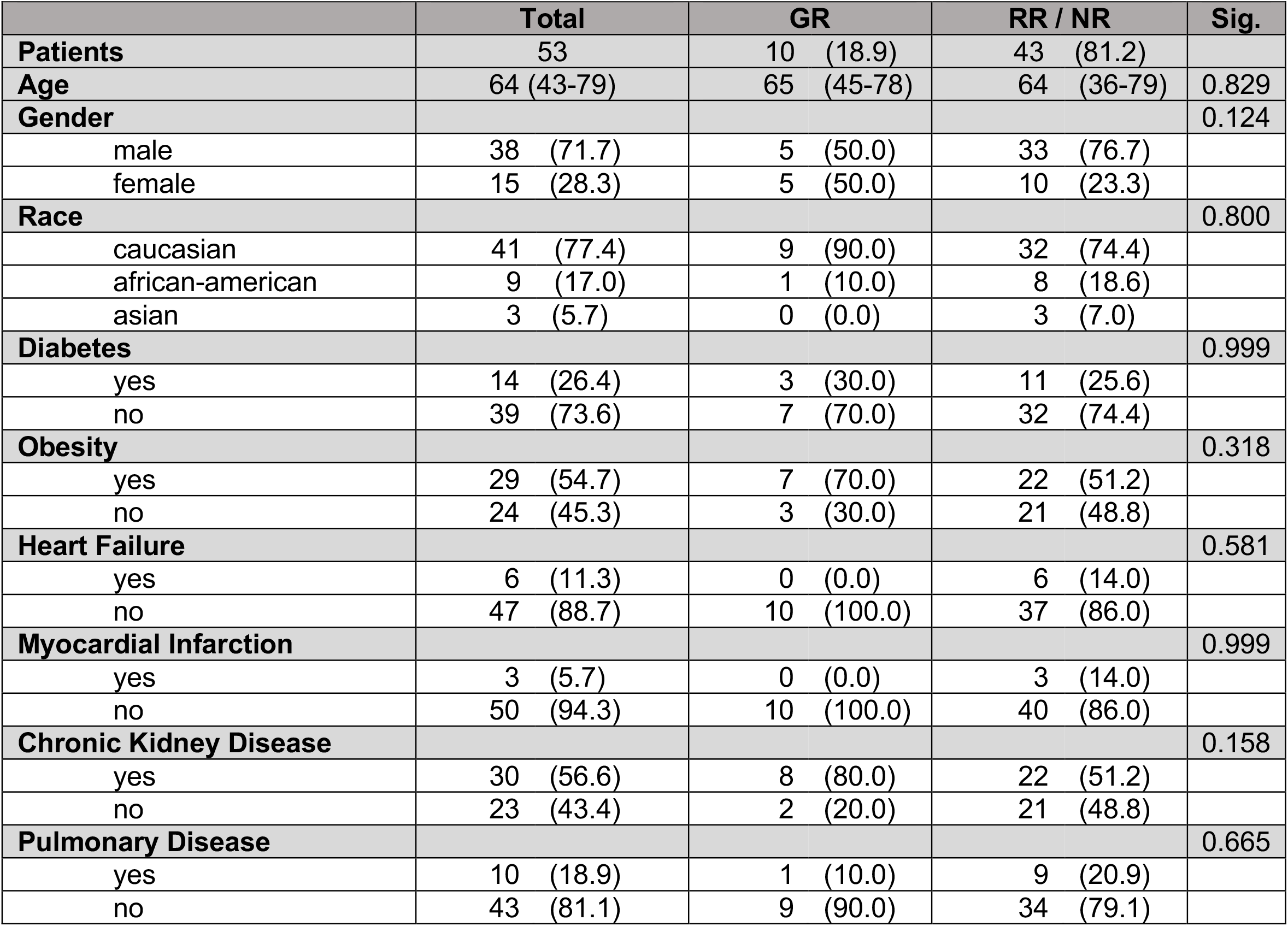
Demographic Characteristics and Medical History.

**Table 2:** Clinical Characteristics at the Time of Vaccination (first dose)

|  | Total | GR | RR / NR | Sig. |
| --- | --- | --- | --- | --- |
| **Patients** | 53 | 10 (18.9) | 43 (81.1) |  |
| **Time from transplant (weeks)** | 322 (30-1260) | 380 (84-698) | 308 (30-1260) | 0.630 |
| **Type of transplant** |  |  |  | 0.175 |
| Kidney | 24 (45.3) | 3 (30.0) | 21 (48.8) |  |
| Liver | 12 (22.6) | 7 (70.0) | 5 (11.6) |  |
| Lung | 6 (11.3) | 0 (0.0) | 6 (14.0) |  |
| Heart | 5 (9.4) | 0 (0.0) | 5 (11.6) |  |
| Kidney / Pancreas | 3 (5.7) | 0 (0.0) | 3 (7.0) |  |
| Heart / Lung | 1 (1.9) | 0 (0.0) | 1 (2.3) |  |
| Liver / Kidney | 1 (1.9) | 0 (0.0) | 1 (2.3) |  |
| Pancreas | 1 (1.9) | 0 (0.0) | 1 (2.3) |  |
| **Induction** |  |  |  | 0.347 |
| Unknown | 3 (5.7) | 0 (0.0) | 3 (7.0) |  |
| Methylprednisolone | 18 (34.0) | 6 (60.0) | 12 (27.9) |  |
| Alemtuzumab | 13 (4.5) | 1 (10.0) | 12 (27.9) |  |
| ATG | 5 (9.4) | 0 (0.0) | 5 (11.6) |  |
| Basiliximab | 6 (11.3) | 1 (10.0) | 5 (11.6) |  |
| Unknown | 8 (15.1) | 2 (20.0) | 6 (14.0) |  |
| **Immunosuppressive Agents (total #)** | 2.0 (1-3) | 1.5 (1-3) | 2.00 (1-3) | 0.018 |
| *Antimetabolite* |  |  |  | 0.007 |
| yes | 38 (71.7) | 3 (30.0) | 35 (81.4) |  |
| no | 15 (28.3) | 7 (70.0) | 8 (18.6) |  |
| *Calcineurin Inhibitor* |  |  |  | 0.647 |
| yes | 48 (90.6) | 10 (100.0) | 38 (88.4) |  |
| no | 5 (9.4) | 0 (0.0) | 5 (11.6) |  |
| *mTOR Inhibitor* |  |  |  | 0.448 |
| yes | 9 (17.0) | 2 (20) | 7 (16.3) |  |
| no | 44 (83.0) | 8 (80) | 36 (83.7) |  |
| *Steroids* |  |  |  | 0.069 |
| yes | 21 (39.6) | 1 (10) | 20 (46.5) |  |
| no | 32 (60.4) | 9 (90) | 23 (53.3) |  |
| **Type of Vaccine** |  |  |  | 0.149 |
| Pfizer | 32 (60.4) | 4 (40.0) | 28 (65.1) |  |
| Moderna | 18 (34.0) | 6 (60.0) | 12 (27.9) |  |
| J&J | 3 (5.7) | 0 (0.0) | 3 (7.0) |  |

### Solid organ transplant recipients maintain their humoral immunity against microbial antigens other than SARS-CoV-2 proteins

To explore whether reduced neutralization was the result of diminished total immunoglobulin levels, we next determined absolute levels of total IgG, IgM, and IgA in the peripheral blood of our patients and controls. We found that the three patient groups (GR, RR, NR) indeed showed significantly lower levels of IgG compared to healthy controls (Figure 2). In contrast, total IgM levels were comparable in patients and controls and total levels of IgA were lower only in the SOTR who had shown a relatively good neutralizing antibody response (Figure 2).

**Figure 2:**
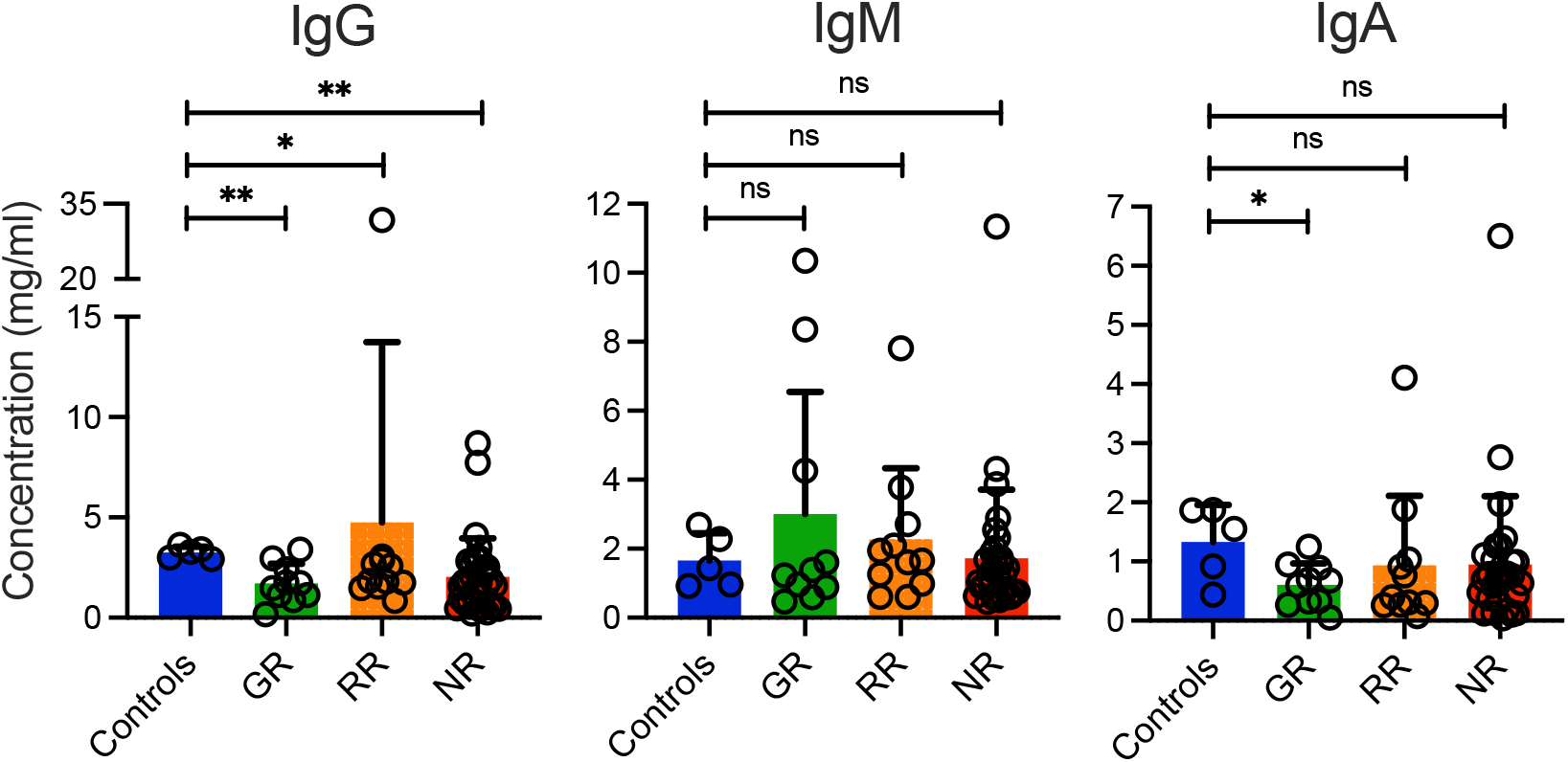
Absolute concentrations of immunoglobulins in the peripheral blood of SOT recipients. Absolute levels of IgG, IgM, and IgA antibodies in our study subjects were measured after the second dose of the vaccine using a commercially available ELISA. Concentrations of total IgG, IgM, and IgA are shown in ng/ml for healthy vaccinated controls and the three different groups of vaccinated SOT patients (Good Responders [GR], Reduced Responders [RR], Non-Responders [NR]) according to the degree of viral neutralization after the second dose of the vaccine. Bars indicate means + SD. Differences between groups were analyzed for statistical significance (*p<0.05, **p<0.01) using the Mann–Whitney U test.

Interestingly, when we determined titers of IgG antibodies against a variety of microbial antigens such as Influenza A H1N1 Nucleoprotein (Flu), tetanus toxoid (TT), Cytomegalovirus Glycoprotein B Protein (CMV), Epstein-Barr virus Glycoprotein gp350 Protein (EBV), and Herpes Simplex Virus Type 1 gD Protein (HSV), we found that NR patients showed even higher antibody titers against Flu, CMV, and EBV. GR patients showed higher antibody levels against CMV compared to healthy controls (Figure 3).

**Figure 3:**
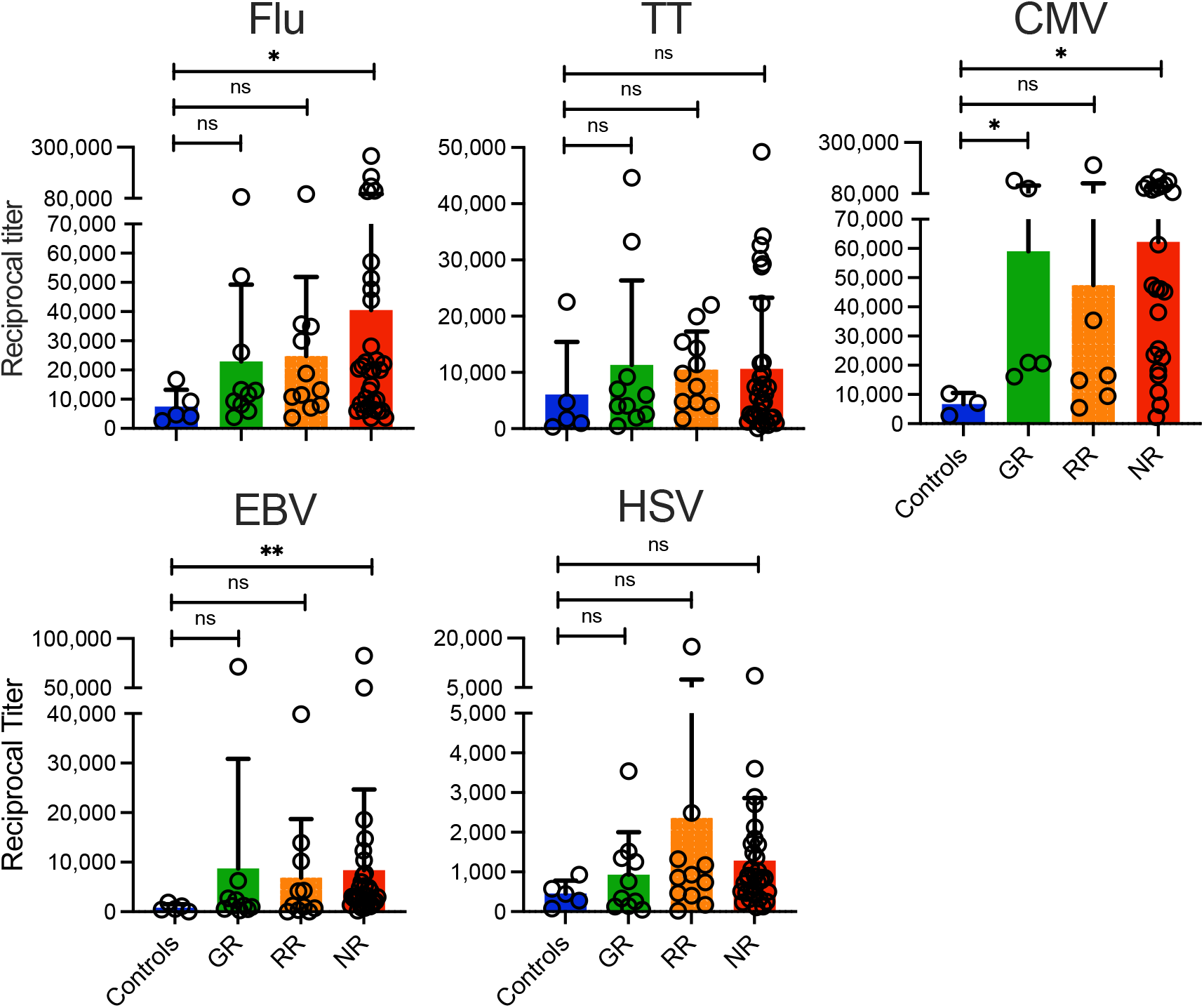
Titers of antibodies against different microbial antigens in the peripheral blood of SOT recipients. Titers of IgG antibodies against full-length recombinant Influenza A nucleoprotein (Flu), tetanus toxoid (TT), cytomegalovirus (CMV), epstein-barr virus (EBV), and Herpes Simplex Virus Type 1 (HSV) were measured in an ELISA. Antibody titers are shown for healthy vaccinated controls and the three different groups of vaccinated SOT patients (Good Responders [GR], Reduced Responders [RR], Non-Responders [NR]) according to the degree of viral neutralization after the second dose of the vaccine. Bars indicate means + SD. Differences between groups were analyzed for statistical significance (*p<0.05, **p<0.01) using the Mann-Whitney U test.

### Vaccine-induced antibody titers against different SARS-CoV-2 proteins are suppressed in solid organ transplant recipients

Next, we measured IgG antibody titers against different SARS-CoV-2 proteins in SOTR post vaccination. We found that, compared to healthy controls, RR and NR patients, and in the case of the delta variant even GR patients, showed significantly lower vaccine-induced antibody titers against the S1 and the RBD proteins as well as their “delta” variants (Figure 4A). In addition, all SOTR showed lower vaccine-induced IgA antibody titers against SARS-CoV-2 proteins S1, RBD and their respective delta variants except for the S2 protein where the GR patients showed antibody levels comparable to healthy donors (Figure 4B). None of the patients or controls showed IgG or IgA antibodies against the SARS-CoV-2 N protein (Figures 4 A+B) indicating that all the antibodies detected were indeed vaccine-induced and not based on a prior COVID-19 infection.

**Figure 4:**
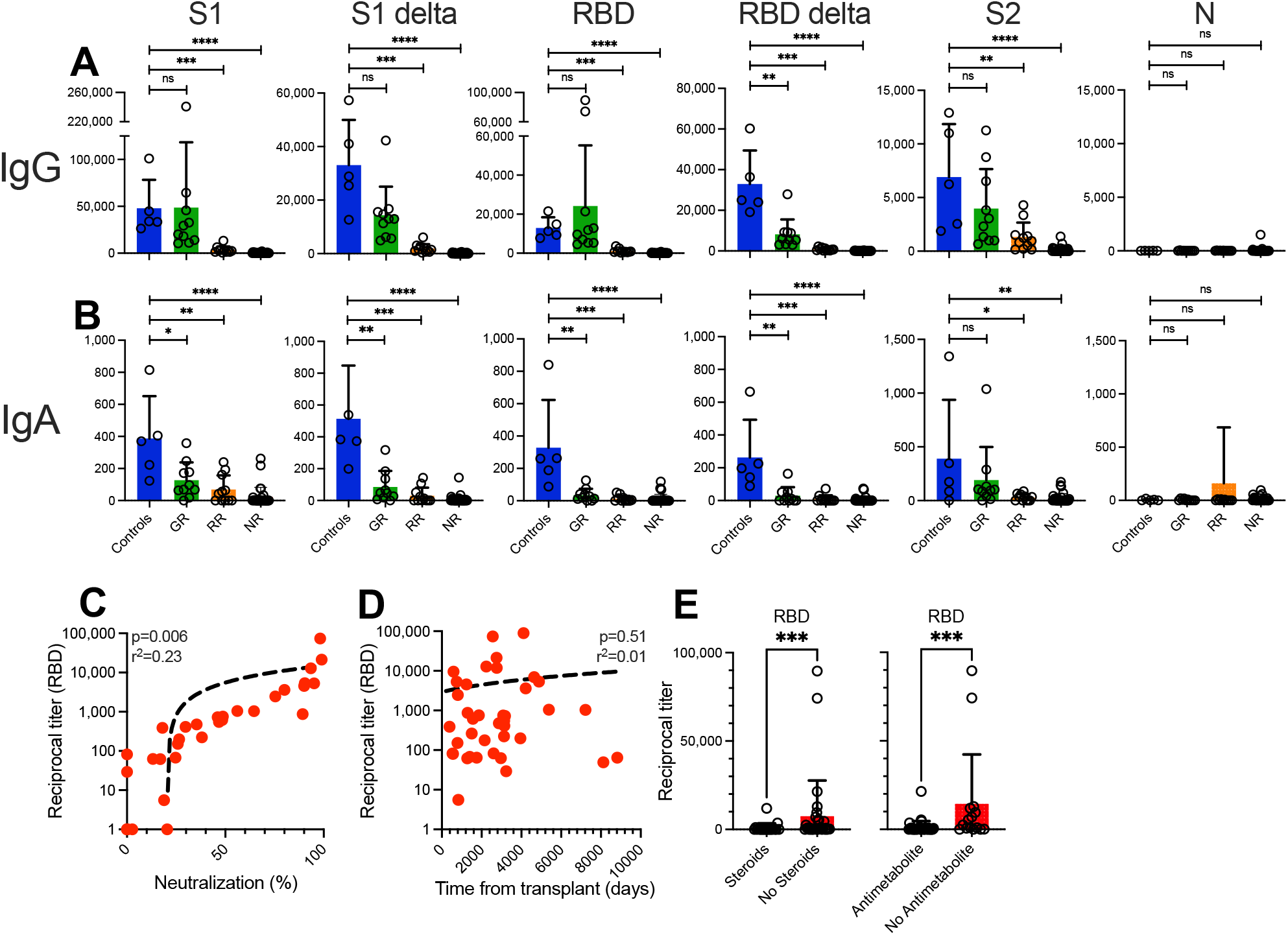
Titers and neutralizing activity anti-SARS-CoV-2 antibodies in SOT recipients after two doses of a COVID-19 mRNA vaccine. Titers of (A) IgG and (B) IgA antibodies against different full-length recombinant SARS-CoV-2 proteins and their delta variants were measured in an ELISA after two doses of a COVID-19 mRNA vaccine. Antibody titers are shown for healthy vaccinated controls and the three different groups of vaccinated SOT patients (Good Responders [GR], Reduced Responders [RR], Non-Responders [NR]) according to the degree of viral neutralization after the second dose of the vaccine. Correlation between anti-RBD IgG antibody titers and (C) neutralizing activity in the same sample and (D) time from SOT at the time of the first dose of the vaccine. (E) Impact of steroid or antimetabolite intake on anti-RBD IgG antibody titers after two doses of the vaccine. Bars indicate means + SD. Differences between groups were analyzed for statistical significance (*p<0.05, **p<0.01, ***p<0.001, ****p<0.0001) using the Mann-Whitney U test.

Overall, these combined data already indicated an association between antibody titers and the neutralizing activity of the patients’ serum. Accordingly, we were able to confirm a highly significant correlation between anti-RBD antibody titers and the neutralizing activity of the patients’ serum after two doses of the vaccine (Figure 4C). All patients with an anti-RBD antibody >4500 showed a viral neutralization of >90%. Analyzing the impact of patient-related characteristics on antibody titers, time from transplant did not have an effect (Figure 4D) titers but intake of steroids or antimetabolites did (Figure 4E).

### Solid organ transplant recipients show a reduced breadth of SARS-CoV-2 vaccine-induced antibody epitopes

Next, we aimed at identifying the most relevant, immunodominant target epitopes of S1 and RBD-specific polyclonal IgG antibodies in vaccinated SOTR when compared to patients with active COVID-19, vaccinated healthy controls (HC), HC before administration of the first dose of the vaccine and non-vaccinated HC whose sera were collected before the COVID-19 pandemic. When we used individual pools of peptides consisting of five 20mer peptides overlapping by 10 amino acids (aa) in an ELISA, we were able to identify regions in the complete S1 protein including its RBD domain that were preferentially targeted by the antiviral antibodies in patients with active COVID-19. As described before [44], for anti-S1 antibodies there were regions within the receptor-binding motif (RBM) corresponding to peptide pool 46-50 (aa 451-510), the C-terminal region of the RBD, and the C-terminal region of the S1 protein adjacent to the RBD that were preferentially targeted (Figure 5). Importantly, HC before administration of the first dose and non-vaccinated HC did not show any measurable responses against any of the S1 protein peptide pools. Although we specifically selected SOTR with the comparably highest antibody titers against the full-length S1 protein, vaccinated SOTR showed a dramatically reduced number of linear B cell epitopes with responses being restricted to a small region adjacent to the C-terminal region of the RBD (Figure 5).

**Figure 5:**
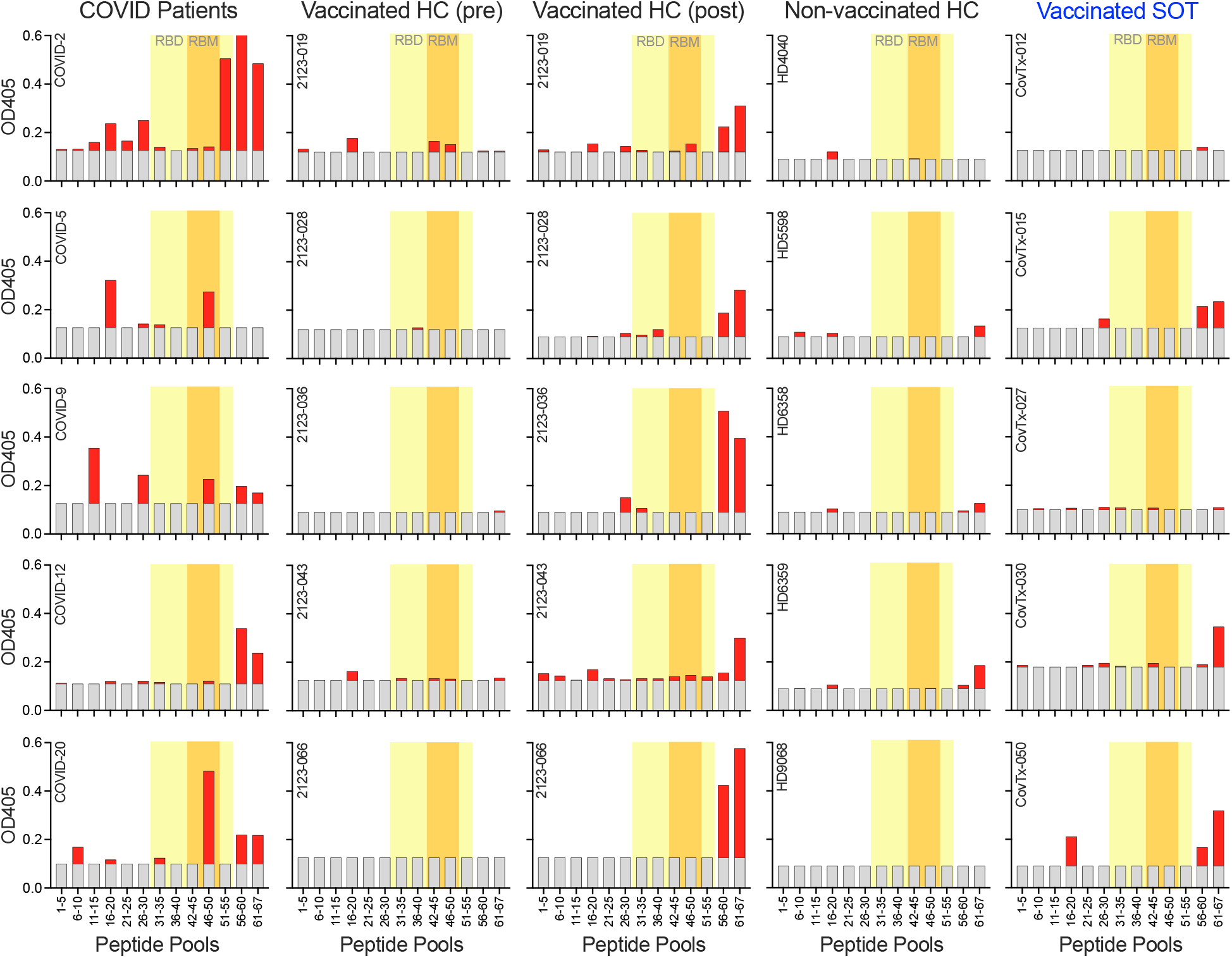
Peptide epitopes within the S1 protein targeted by vaccine-induced antibodies. Plasma samples from 5 COVID-19 patients, 5 vaccinated healthy controls after two doses of the vaccine, the same controls before receiving the first dose of the vaccine, 5 non-vaccinated healthy controls, and 5 SOT recipients with known anti-SARS-CoV-2 reactivity after the second dose of the vaccine were analyzed for immunodominant peptide epitopes. Peptide pools of 5 20mer peptides each overlapping by 10aa were used in an ELISA. Gray bars indicate background levels. RBD and RBM regions within the S1 protein are highlighted in yellow and orange, respectively.

### In solid organ transplant recipients, an additional vaccine dose results in a dramatically improved antibody responses against all SARS-CoV-2 variants except omicron

Next, we analyzed antibody responses induced by an additional vaccine dose in our SOTR. We found that an additional dose led to a highly significant increase in antibody titers against SARS-CoV-2 proteins RBD, S1, and S2 (Figure 6A). Importantly, the additional dose did not only lead to an increase in IgG antibody titers but also an enhanced neutralizing activity of the polyclonal sera. Among the 32 SOTR that received an additional vaccine dose, numbers of GR (neutralizing activity >90%) increased from 6/32 (18.8%) after the second vaccine dose to 18/32 (56.3%) after the additional vaccine dose. The number of NR patients with no (<30%) neutralizing activity decreased from 15/32 (46.9%) to as few as 7/32 (21.9%) when the wild-type RBD protein was used (Figure 6B). The improvement in neutralizing activity was even more pronounced when we used the delta and “UK” (alpha) variants of the RBD protein (Figure 6B). Numbers of GR for both variants increased from as few as 2/32 (0.6%) to 19/32 (59.4%) and numbers of NR patients decreased from 18/32 (56.3%) to 8/32 (25%) and from 21/32 (65.6%) to 7/32 (21.9%), respectively (Figure 7B). For the beta variant there was not a single SOTR showing a neutralization >90% after just two vaccine doses. While the rate increased to 13/32 (40.6%) after the third dose, the proportion of “post-third dose” GRs was still substantially lower for the beta variant than for the aforementioned variants and the wild-type RBD (Figure 6B). Interestingly, when we analyzed patients not converting vs. converting from NR to RR/GR following an additional vaccine dose we found no significant differences with regard to antimetabolite intake (5/7 vs. 7/8), or time between booster vaccination and blood sampling (37 [14-66] vs. 62 [18-186] days).

**Figure 6:**
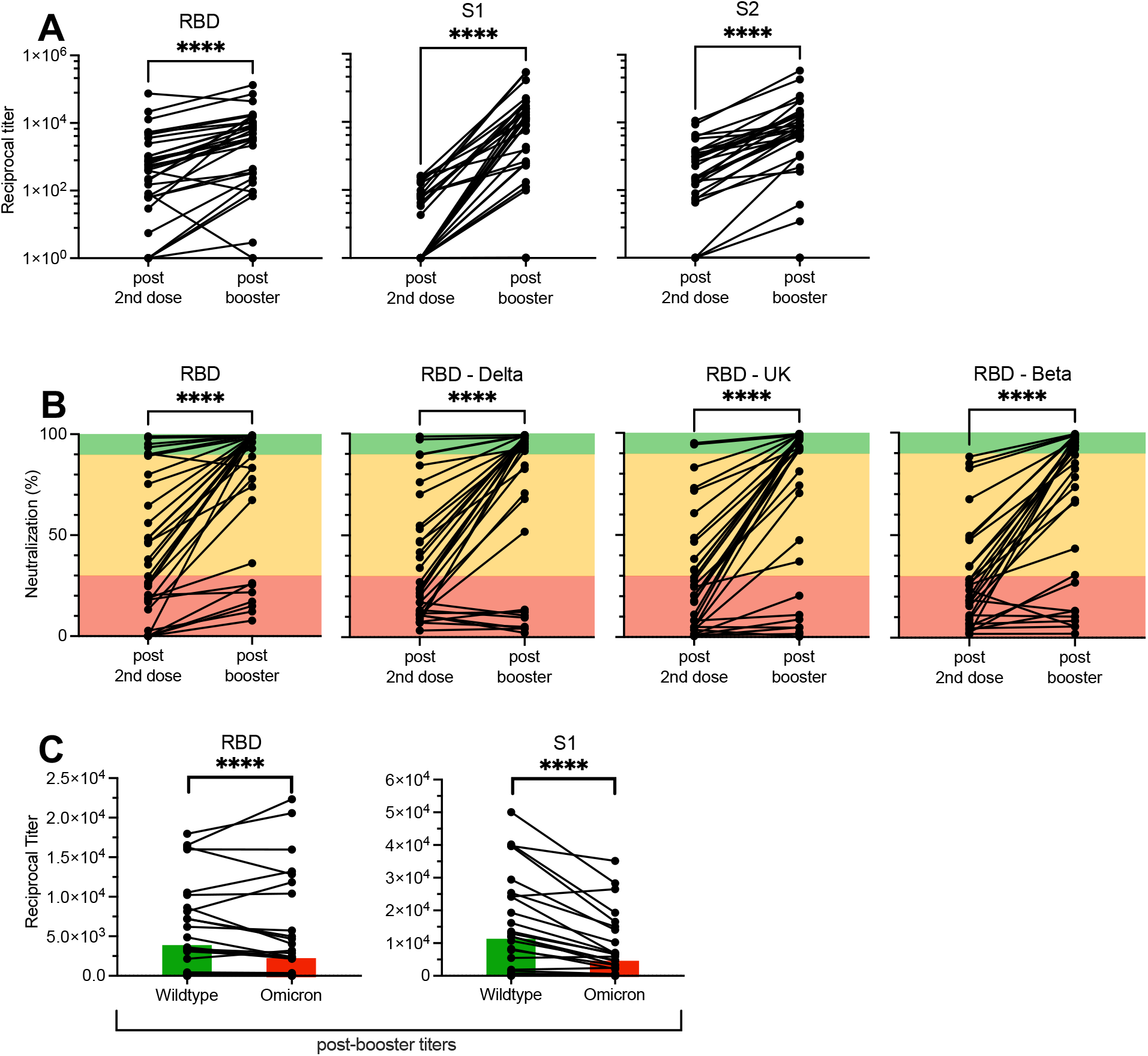
Effect of a third dose of an mRNA vaccine on anti-SARS-CoV-2 antibodies and neutralizing activity. (A) Titers of IgG antibodies against different full-length recombinant SARS-CoV-2 proteins and their delta variants were measured in 32 SOT recipients using an ELISA before and after a third “booster” dose of a COVID-19 mRNA vaccine. (B) Neutralizing activity before and after a third “booster” dose of a COVID-19 mRNA vaccine in the peripheral blood of the same SOT recipients. Green, orange, and red areas indicate different degrees of inhibition (green: >90%, orange: 30-89%, red: <30%). (C) Titers of post-booster IgG antibodies against the original anti-SARS-CoV-2 RBD and S1 proteins vs. their omicron variants. Bars indicate median levels. Differences between groups were analyzed for statistical significance (*p<0.05, **p<0.01, ***p<0.001, ****p<0.0001) using the Mann-Whitney U test.

Finally, we compared binding of our patients’ post-third dose anti-RBD and anti-S1 IgG antibodies to the respective wild-type SARS-CoV-2 proteins vs. their omicron variants. We observed a highly significant decrease by 30% and 58% in median anti-RBD and anti-S1 antibody titers, respectively, when these were exposed to the omicron variants vs. the wild-type proteins (Figure 6C).

## DISCUSSION

Here, we present the results of our prospective study investigating humoral immune responses to multiple SARS-CoV-2 viral variants following initial vaccination with COVID-19 vaccines and following an additional vaccine dose in SOTR. Our analyses show that most SOTR do not respond adequately to the initial vaccine series. The majority of our vaccinated SOTR were “Non-Responders” with a viral neutralization of less than 30%. Less than 20% of SOTR showed a neutralizing activity comparable to our healthy vaccinated controls. Few studies have assessed neutralizing activity in SOTR after mRNA vaccination with one study demonstrating, in line with our results, a neutralizing activity of >30% in only 27% of all patients [47]. In conclusion, only a relatively small subgroup of SOTR is able to generate functionally relevant antibodies after two doses of a COVID-19 mRNA vaccine.

When we asked whether the reduced immunoreactivity of our SOTR was an expression of a broadly suppressed humoral immunity, we found that absolute IgG levels in the patients’ peripheral blood at the time of the first dose of the vaccine were indeed lower than in healthy controls. However, titers of IgG antibodies against a variety of microbial antigens such as Influenza, CMV, and EBV were even higher in the SOTR, especially the ones with a reduced anti-SARS-CoV-2 response, than in healthy controls. These data indicate that humoral responses against recall antigens are not substantially impaired in SOTR but that the problem is a dysfunctional priming of antibody-mediated immune response against novel antigens such as the S protein of SARS-CoV-2 that the patient has never encountered before, possibly based on impaired T cell help during priming and/or limited B cell expansion.

Analyzing IgG antibody titers against different SARS-CoV-2 proteins in SOTR we found that especially patients with a reduced anti-SARS-CoV-2 neutralizing activity showed significantly reduced vaccine-induced antibody titers against the S1 and the RBD proteins of the virus including their delta variants. Importantly, anti-SARS-CoV-2 antibody titers correlated significantly with the neutralizing activity of the patients’ serum after two doses of the vaccine and all patients with an anti-RBD antibody >4500 showed a viral neutralization of >90%, supporting the functional relevance of measuring antibody titers in vaccinated patients, especially those with an impaired humoral immune response.

Out of a large number of clinical variables, we only found the overall intensity of the immunosuppressive treatment to be associated with a reduced vaccine-induced SARS-CoV-2 neutralizing activity and reduced levels of anti-RBD antibodies. Specifically, treatment with antimetabolites had the most significant negative impact on the patients’ antiviral humoral immunity. This finding is in agreement with recent studies by other groups showing that SOTR on treatment with antimetabolites exhibit a reduced response to two doses of mRNA-based SARS-CoV-2 vaccines [44, 48-51]. Interestingly, it has previously been shown that SOTR show suboptimal responses to seasonal influenza vaccination with seroconversion rates as low as 34% and lower seroconversion rates were associated with high doses of MMF and lower numbers of postvaccine H1N1-specific IL-4^+^CD4^+^ T cells [52].

Mycophenolic acid (MPA) is a selective, non-competitive, and reversible inhibitor of inosine-50-monophosphate dehydrogenase (IMPDH). MPA inhibits the production of guanosine and deoxyguanosine nucleotides leading to a reduced proliferation of T and B lymphocytes and impaired production of immunoglobulins through the guanosine and deoxyguanosine nucleotides in depleted lymphocytes [53]. Mycophenolate mofetil (MMF) is a prodrug of MPA and suppresses T cell responses to allogeneic cells and other antigens. Importantly, *in vivo* the drug also suppresses primary, but not secondary, antibody responses [54]. In one study, patients with systemic lupus erythematosus (SLE) taking MMF showed lower numbers of plasmablasts [55], a finding that was later confirmed by two different groups [56, 57]. Interestingly, another study investigating B cell responses to the SARS-CoV-2 BNT162b2 vaccine in kidney transplant recipients also observed diminished seroconversion rates correlating with a reduced generation of plasmablasts and memory B cells [40]. Therefore, future studies should evaluate in detail the effect of MMF intake on B cell and T cell phenotype and function after COVID-19 vaccination to help improve immune responses in SOTR.

Even though we specifically selected SOTR with the comparably highest anti-S1 antibody titers, vaccinated SOTR showed a dramatically reduced number of linear B cell epitopes, indicating an overall reduced functional B cell diversity in these patients. Vaccine-induced antibody responses against linear peptides in SOTR were primarily directed against a small region adjacent to the C-terminal region of the RBD, a region that we and others have previously described as immunodominant [44, 58]. Future studies should investigate whether the reduced breadth of the humoral immune response in SOTR further contributes to the immune escape of the SARS-CoV-2 virus in addition to lower overall antibody titers.

Next, we analyzed whether an additional dose ofSARS-CoV-2 vaccine in our SOTR could improve humoral responses observed after completion of the initial vaccine series. Importantly, we found that an additional dose not only led to a marked increase in antibody titers against the different SARS-CoV-2 proteins but also enhanced neutralizing activity of the polyclonal sera. Remarkably, the increase in neutralizing activity after the additional vaccine dose was even more pronounced for the delta and the alpha / UK variants of the RBD protein. In contrast, the beta variant, which shares immunorelevant mutations with the omicron variant, showed less improvement when compared to the aforementioned variants.

The omicron (B.1.1.529) variant of SARS-CoV-2 was only very recently detected in South Africa and due to enhanced transmissibility it has rapidly spread and has become the dominant variant in many countries, including the United States [59]. A hallmark of the omicron variant is the large number of S protein mutations potentially causing immune escape even in fully vaccinated healthy individuals [43]. Here, we compared binding of our patients’ anti-RBD and anti-S1 IgG antibodies following an additional vaccine dose to the respective wild-type SARS-CoV-2 proteins vs. their omicron variants. We observed a highly significant decrease in median anti-RBD and anti-S1 antibody titers, respectively, when these were exposed to the omicron variants. Given the strong correlation between anti-RBD/S1 antibody titers and neutralizing activity demonstrated above, we consider it highly likely that this decreased antibody binding is of functional and possibly clinical relevance. We think that findings like the ones presented herein have the potential to improve currently available prophylactic approaches for SOTR, e.g. the use of additional vaccine doses or other types of pre- or post-exposure prophylaxis, all of which may be particularly important given the recent occurrence of immune escape variants of SARS-CoV-2. In this context we would like to stress that multi-mutational variants of the virus are much more likely to arise in immunocompromised patients with a prolonged course of the COVID-19 infection [60]. This is another important reason to find ways to prevent these individuals from becoming infected or at least limit the duration of viral persistence in case of an active COVID-19 infection.

## Data Availability

All data produced in the present study are available upon reasonable request to the authors.

## DECLARATIONS

### Authors’ contributions

K.K.S., J.S.H., S.V.N., S.D., and A.P.R. designed the study, collected patient samples, analyzed the data, and wrote the manuscript. K.S. collected patient samples, analyzed the data, and wrote the manuscript. Y.J.Y, P.L., H.D.M., A.C., W.Y.X., A.C., and J.A. collected patient samples. M.M. and A.A. collected patient data. T.I., X.F., D.O., and S.V.A. performed experiments and wrote the manuscript. T.L., N.M.H., E.V.M, K.H., J.B., and O.G. analyzed the data and wrote the manuscript. D.A. designed the study, performed experiments, analyzed the data, made figures, and wrote the manuscript.

## ACKNOWLEDGEMENTS

We would like to thank Natalie McNally for coordinating patient enrollment and Marion Williams for sample collection. This study was funded by two grants from the Kahlert Foundation. In addition, the study was supported in part by funds from the Division of Infectious Diseases and the Transplantation Program.

