## Supplementary figures and images for "Humoral immune responses against SARS-CoV-2 variants including omicron in solid organ transplant recipients after three doses of a COVID-19 mRNA vaccine"

### Supplemental Figures

#### Slide 1
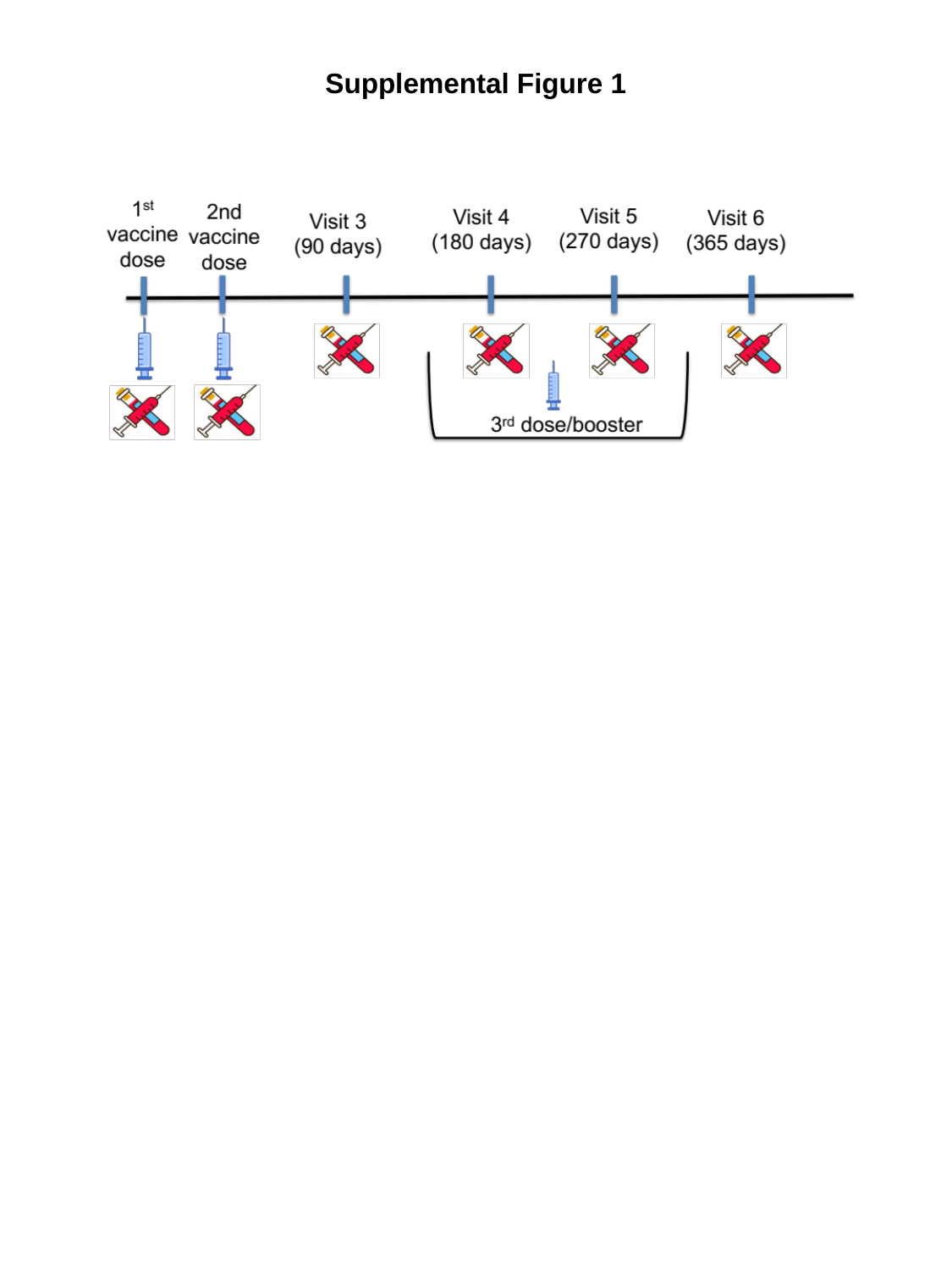

Supplemental Figure 1
