## Supplemental Tables for "Humoral immune responses against SARS-CoV-2 variants including omicron in solid organ transplant recipients after three doses of a COVID-19 mRNA vaccine"

**Supplemental Table 1: Control Proteins**

| **Protein** | **Expression System** | **Manufacturer** | **Catalogue No.** |
| --- | --- | --- | --- |
| Influenza A H1N1 Nucleoprotein (NP) | Baculovirus-Insect Cell | Sino Biological | 11675-V08B |
| Epstein-Barr Virus (EBV) Glycoprotein gp350 | Baculovirus-Insect Cell | Sino Biological | 40373-V08B |
| Cytomegalovirus (CMV) Glycoprotein B | HEK293 | Sino Biological | 10202-V08H1 |
| Glutathione S-Transferase Alpha 1 (GSTA1) | HEK293 | Sino Biological | 15237-H08H |
| Herpes Simplex Virus (HSV) Type 1 gD | E. coli | Abcam | ab43045 |
| Tetanus Toxoid |  | Calbiochem | 582231 |

**Supplemental Table 2: SARS-CoV-2 Proteins**

| **Protein** | **Variant** | **Mutations** | **Expression System** | **Conjugate** | **Manufacturer** | **Catalogue No.** |
| --- | --- | --- | --- | --- | --- | --- |
| Spike S1 |  |  | HEK293 |  | Acro Biosystems | S1N-C52H2 |
| Spike Receptor-Binding Domain (RBD) |  |  | HEK293 |  | Acro Biosystems | SPD-C52H2 |
| Spike S2 |  |  | HEK293 |  | Acro Biosystems | S2N-C52H5 |
| Nucleocapsid (N) |  |  | HEK293 |  | Acro Biosystems | NUN-C5221 |
| Spike S1 | Delta (B.1.617.2) | T19R, G142D, EF156-157del, R158G, L452R, T478K, D614G, P681R | HEK293 |  | Acro Biosystems | S1N-C52Hu |
| Spike Receptor-Binding Domain (RBD) | Delta (B.1.617.2) | L452R, T478K | HEK293 |  | Acro Biosystems | SPD-C52Hh |
| Spike S1 | Omicron (B.1.1.529) | A67V, HV69-70del, T95I, G142D, VYY143-145del, N211del, L212I, ins214EPE, G339D, S371L, S373P, S375F, K417N, N440K, G446S, S477N, T478K, E484A, Q493R, G496S, Q498R, N501Y, Y505H, T547K, D614G, H655Y, N679K, P681H) | HEK293 |  | Acro Biosystems | S1N-C52Ha |
| Spike Receptor-Binding Domain (RBD) | Omicron (B.1.1.529) | G339D, S371L, S373P, S375F, K417N, N440K, G446S, S477N, T478K, E484A, Q493R, G496S, Q498R, N501Y, Y505H | HEK293 |  | Acro Biosystems | SPD-C522e |
| Receptor-Binding Domain (RBD) | Alpha / UK (B.1.1.7) | N501Y | HEK293 | HRP | GenScript | Z03595 |
| Receptor-Binding Domain (RBD) | Beta (B.1.351) | K417N, E484K, N501Y | HEK293 | HRP | GenScript | Z03596 |
| Receptor-Binding Domain (RBD) | Delta (B.1.617) | L452R, T478K | HEK293 | HRP | GenScript | Z03614 |
| Receptor-Binding Domain (RBD) | Delta Plus (B.1.617.2.1) | K417N, L452R, T478K | HEK293 | HRP | GenScript | Z03690 |
